# Evaluating the impact of unadjusted confounding and study design on estimated pathogen-attributable incidence of diarrhoea among children in low and middle-income countries: a sensitivity analysis of an attribution algorithm in the MAL-ED cohort

**DOI:** 10.1101/2025.06.24.25330206

**Authors:** Charlotte Doran, James Platts-Mills, Eric Houpt, Jie Liu, Elizabeth Rogawski McQuade

**Affiliations:** Department of Epidemiology, Emory University Rollins School of Public Health; Division of Infectious Diseases and International Health, University of Virginia; School of Public Health, Qingdao University, Qingdao, China

## Abstract

**Background:** Understanding diarrhoea aetiology is critical for understanding vaccine impact, but causal attribution is difficult. We evaluated the sensitivity of an existing attributable fraction-based method to unadjusted confounding and study design.

**Methods:** We used MAL-ED data to estimate attributable incidence (IR_attr_) using an algorithm regressing diarrhoea incidence on pathogen quantity. The reference model used mixed-effects logistic regression adjusted for co-infection, sex, test batch, and age. We evaluated the magnitude of confounding by prior immunity, antibiotics, socioeconomic status, and breastfeeding in incrementally-adjusted models. To understand the impact of post-diarrheal shedding, we excluded stools collected ±7, 14, or 28 days from an episode. We conducted matched nested case-control and case-crossover studies and used Poisson regression to calculate risk-based estimates of the IR_attr_.

**Results:** 40 406 stools samples from 1715 children were included (6625 diarrhoeal, 33 781 control). Prior immunity had the greatest confounding impact, with adjusted models underestimating IR_attr_ compared to the reference; overall, confidence intervals overlapped substantially. Excluding samples proximal episodes showed no consistent pattern of bias, and most estimates were ±10% of the reference except Cryptosporidium, tEPEC, and norovirus. Nested case-control and case-crossover study designs produced similar IR_attr_ estimates, and risk-based estimates were lower than odds-based estimates, ranging from 33% (Shigella) to 49% (tEPEC) fewer episodes per 100 child-years compared to the reference.

**Conclusion:** This algorithm was robust to unadjusted confounding, but risk- and odds-based estimates differed. We recommend adjusting for breastfeeding, antibiotics use, and prior infection using risk-based model. If case-control design is necessary, matched incidence density sampling should be used.

## Introduction

Despite reductions in incidence in recent decades, diarrhoeal disease remains a leading cause of morbidity and mortality in children under two, accounting for 6.1% of deaths among infants <1 year and 13.6% of deaths among children 12-23 months in 2021(1,2). Diarrhoea burden is especially high in low- and middle-income countries, causing 15% of deaths among children 12-23 months in Sub-Saharan Africa and South Asia(1). Rotavirus vaccination programs have reduced the incidence of severe disease and hospitalization in children(3), but vaccine development for other enteropathogens is slow(4,5).

Key to understanding the potential impact of novel vaccines is accurate estimation of aetiology-specific diarrhoea burden. This estimation is non-trivial, and results vary considerably depending on study design and method(6). Some studies interrogating aetiology used hospital-based cases and considered episodes as caused by a pathogen if that pathogen was detected (7–9). Detection of multiple pathogens during episodes is common due to prolonged shedding and subclinical infections(10); therefore, detection alone is insufficient to determine aetiology. We previously described an attributable fraction (AF)-based algorithm for estimation of attributable incidence using data from the longitudinal community-based MAL-ED) birth cohort(11). This method included prospective collection of diarrheal and non-diarrheal stools and used quantitative molecular diagnostics to estimate the association between pathogen quantity and diarrhoea, which was used to calculate each pathogen’s population AF.

Using quantitative methods to identify cases of diarrhoea is preferable to detection alone because higher pathogen quantity has a stronger association with diarrhoea than low but detectable levels for most pathogens(12). However, estimates necessarily come from observational studies, often case-control or case-only studies, which can be subject to confounding and selection bias. Prior infection and breastfeeding convey immunity and reduce the risk of future infection and diarrhoea in infants and therefore may be important confounders(4,13–15). Similarly, widespread use of antibiotics in high-burden settings may lower pathogen quantity and confound its association with diarrhoea, and prolonged shedding following infection may lead to misclassification of the outcome as causally related to the pathogen of interest. Finally, odds-based estimates of the AF rely on the rare disease assumption; meaning, the odds ratio will approximate the risk ratio only when the outcome is rare. In high-burden settings such as MAL-ED, odds-based estimates may overstate aetiology-specific burden of diarrhoea compared to risk-based estimates. We aim to extend the AF method to account for these biases and evaluate its sensitivity to unadjusted confounding by antibiotics use, breastfeeding, and other demographic factors, study definitions of diarrhoeal episodes, and study design.

## Methods

### Study Participants

The MAL-ED study design has been previously described(16). Between November 2009 and February 2012, singleton children born of mothers aged ≥16 years were enrolled ±17 days of birth in eight study locations: Dhaka, Bangladesh; Fortaleza, Brazil; Vellore, India; Bhaktapur, Nepal; Naushero Feroze, Pakistan; Loreto, Peru; Venda, South Africa; and Heydom, Tanzania. Children were included if their family intended to remain in the study area for ≥6 months from enrolment and they had no siblings enrolled in the study. Children with a birthweight or enrolment weight of less than 1500 g or who were diagnosed with a congenital disease, severe neonatal disease, or enteropathy were excluded. All sites received ethics approval from their respective governmental, local institutional, and collaborating institutional ethics review boards. Study staff obtained written consent from the parent or guardian of every child.

### Data and Sample Collection

At enrolment, study staff recorded each child’s sex, birthdate, and information about breastfeeding and household characteristics. Fieldworkers visited the families of enrolled children twice weekly for diarrhoea and antibiotics use surveillance via a standardized questionnaire. We defined diarrhoea as maternal report of ≥3 loose stools within 24 hours or one loose stool with visible blood. Any episodes separated by ≥48 hours without diarrhoea were considered distinct. Fieldworkers collected stool samples during diarrhoeal episodes (case stools) and during monthly home visits without diarrhoea (control stools). Caregivers reported the duration of exclusive breastfeeding during twice-weekly and monthly home visits for the first 8 months and monthly thereafter. Stool samples were analysed by a standardized protocol as previously described(11,17,18). The pathogens with the highest estimated attributable incidence were included in the present analysis: adenovirus 40/41, astrovirus, *Campylobacter jejuni* or *Campylobacter coli*, *Cryptosporidium*, norovirus GI and GII (modelled as combined), rotavirus, sapovirus, *Shigella*, typical enteropathogenic *Escherichia coli* (tEPEC), and enterotoxigenic *Escherichia coli* (heat-stable [ST ETEC] and heat-labile [LT ETEC]; modelled as combined).

### Statistical Analysis

#### Calculation of attributable incidence

The reference model estimating attributable incidence rates (IR_attr_) has been previously described(11). We used the quantification cycle value as an inverse measure of pathogen quantity, with a value of 35 indicating no detection. We estimated pathogen-specific burden using Afs calculated with pathogen prevalence and the association of pathogen quantity and diarrhea(19). In the reference model, we used a generalized linear mixed-effects model (GLMM) regressing diarrhoea on pathogen quantity, adjusted for quantity of other pathogens, sex, test batch, age (days, linear and quadratic), a random slope for site, and a random intercept for each individual. All samples were included in the reference analysis. We first calculated population AF using the episode and pathogen quantity-specific odds ratios from the GLMM, defined as the ratio of predicted odds of diarrhoea using the observed pathogen quantity versus no detection, and summing over each episode, *i*:

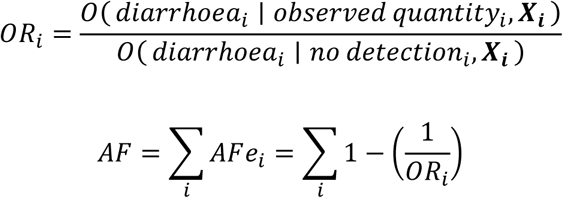

Where ***X*** is the set of predictors in the GLMM. We then calculated IR_attr_ as the product of the total number of diarrhoea episodes, *d*, and the AF divided by the number of child years at risk.

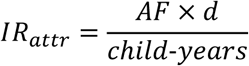

We estimated 95% confidence intervals (CIs) using bootstrapping with 1000 iterations. Analyses were conducted using R version 4.4.1 with the “lme4”, “nloptr”, and “ccoptimalmatch” packages.

#### Sensitivity on potential confounders

We selected potential confounders *a priori* using a directed acyclic graph (DAG, Error! Reference source not found.) informed by literature review, with only strongly plausible causal relationships retained. Age and prior exposure to pathogens have been shown to independently impact diarrhoea incidence in children(20,21). We modelled prior exposure as categorical with levels of zero, one, two, and three or more prior immunizing exposures to the pathogen of interest, defined as having any stool samples with the pathogen detected at a level with the potential to confer natural immunity(22). Breastfeeding was defined as the proportion of the 30 days prior to sample collection in which the child was exclusively breastfed. Antibiotic use, limited to macrolides and fluoroquinolones, was defined in two etiologically relevant periods of use: 1-15 days prior and 16-30 days prior to the stool sample. Socioeconomic status (SES) was measured using the WAMI index, developed and validated for MAL-ED as a standardized measure of SES. The WAMI index is a composite score (0–1) of improved <u>w</u>ater and sanitation, <u>a</u>ssets, <u>m</u>aternal education, and household income, where a higher score indicates a higher SES(23). Because all case stools were processed in the same batch, test batch was included as a study-specific confounder. Covariates were evaluated individually and in a fully adjusted model containing to assess the magnitude of unadjusted confounding.

#### Sensitivity on sample inclusion

To determine the impacts of inclusion of control samples collected proximal to a diarrhoeal episode, we conducted a sensitivity analysis on inclusion criteria with three levels, in which samples that were collected ±7, 14, or 28 days of a case sample were excluded from the analysis. If two or more diarrhoea episodes occurred with less than the specified timeframe between consecutive episodes, only the sample from the earliest episode was retained. We used the reference model for all sensitivity analyses on sample inclusion criteria.

#### Sensitivity on study design

We conducted three sensitivity analyses on study design. First, we matched case samples one-to-one to control samples from a different child on sex, site, and age ±14 days in a nested case-control analysis, maximizing the proportion of case samples that could be matched to control samples while minimizing residual confounding by age. The same child could serve as a case and a control more than once, but case samples could not be matched to control samples from the same child. Second, we used a case-crossover design, matching case samples one-to-one to control samples collected between 4 and 60 days prior from the same child(24). Non-diarrheal stools that occurred after the case stool sample were ineligible for selection as controls due to prolonged viral shedding after symptoms resolve(10,25). Finally, we calculated risk-based IR_attr_ estimates with the full sample using Poisson regression with a log link. Covariates for the Poisson model were identical to the reference model. The predicted risks were used to calculate the episode-specific risk ratio and population AF, where ***X*** is the set of predictors in the GLMM.

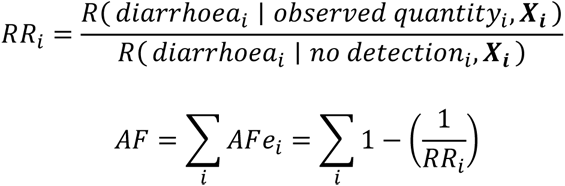

Bootstrapped 95% CIs were resampled at the matching stratum level for the nested case-control and nested-case crossover design.

## Results

In total, 1715 children were included in the analysis, contributing 44 570 stool samples over 41 123 child-months. Of those samples, 40 406 had sufficient specimens for testing and valid qPCR results (6625 case stools and 33 781 control stools). Cohort characteristics have been previously described(11). The two-year risk of infection was highest for sapovirus (77%) and norovirus GII (75%) (**Table 1**). Use of antibiotics was more common prior to diarrhoeal stools than control stools, with 17.0% use within 15 days prior to diarrhoeal stools and 5.0% use prior to control stools. Only 15.2% of stool samples had any exclusive breastfeeding within 30 days prior. Cases were more likely to have prior exposure to pathogens than controls, and, among children <6 months old, case stools were less likely to have exclusive breastfeeding compared to control stools (**Table 2**).

**Table 1.**
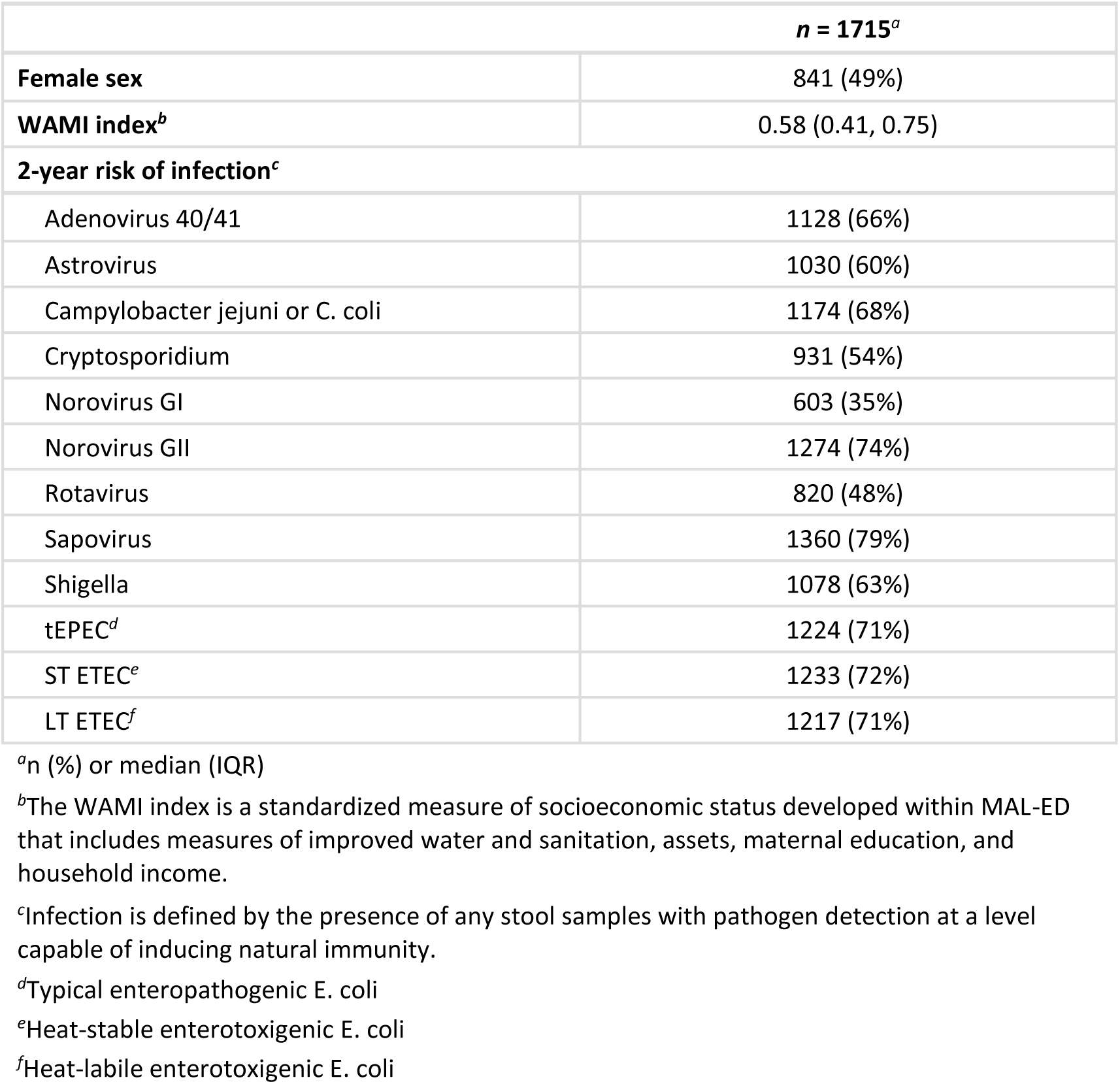
Characteristics and infection risk of children included in the MAL-ED cohort.

**Table 2.** Stool sample-level characteristics for monthly surveillance (non-diarrheal) and diarrheal stools in the MAL-ED cohort.

|  | Non-Diarrheal<br>*n* = 33 781<sup>a</sup> | Diarrheal<br>*n* = 6625<sup>a</sup> |
| --- | --- | --- |
| **Test batch** |  |  |
| First batch | 13 884 (41%) | 6625 (100%) |
| Second batch | 19 897 (59%) | 0 (0%) |
| **Antibiotics use (0-15 days prior)** | 1683 (5.0%) | 1111 (17%) |
| **Antibiotics use (16-30 days prior)** | 1581 (4.7%) | 634 (9.6%) |
| **Proportion of last 30 days exclusively breastfed** | 0.00 (0.00, 0.00) | 0.00 (0.00, 0.00) |
| <6 months old | 0.33 (0.00, 1.00) | 0.10 (0.00, 0.75) |
| **Any prior immunity<sup>b</sup>** |  |  |
| Adenovirus 40/41 | 13 598 (40%) | 3318 (50%) |
| Astrovirus | 12 594 (37%) | 3259 (49%) |
| Campylobacter jejuni or C. coli | 16 090 (48%) | 3528 (53%) |
| Cryptosporidium | 8400 (25%) | 1807 (27%) |
| Norovirus GI | 6451 (19%) | 1559 (24%) |
| Norovirus GII | 17 681 (52%) | 4103 (62%) |
| Rotavirus | 10 452 (31%) | 2784 (42%) |
| Sapovirus | 15 867 (47%) | 3665 (55%) |
| Shigella | 9380 (28%) | 1888 (28%) |
| tEPEC<sup>c</sup> | 15 925 (47%) | 3439 (52%) |
| ST ETEC<sup>d</sup> | 14 617 (43%) | 3112 (47%) |
| LT ETEC<sup>e</sup> | 14 967 (44%) | 3106 (47%) |

<sup>a</sup>n (%); Median (Q1, Q3)
<sup>b</sup>Prior immunity is defined as having any prior stool samples with a pathogen quantity detected at a level capable of conferring natural immunity.
<sup>c</sup>Typical enteropathogenic E. coli
<sup>d</sup>Heat-stable enterotoxigenic E. coli
<sup>e</sup>Heat-labile enterotoxigenic E. coli

Using the reference model, rotavirus (IR_attr_ = 25.0 [22.5 – 28.7]), adenovirus 40/41 (IR_attr_ = 20.8 [17.7 – 24.9]), sapovirus (IR_attr_ = 19.9 [16.4 – 24.1]), and ETEC (IR_attr_ = 17.2 [14.6 – 21.9]) had the highest attributable incidence in the first year of life. In the second year of life, *Shigella* was the leading cause of diarrhoea (IR_attr_ = 41.6 [38.3 – 47.2]), and sapovirus (IR_attr_ = 24.9 [21.0 – 29.6]) and ETEC (IR_attr_ = 20.4 [17.2 – 25.7]) incidence also increased from the first year (**Figure 1**). These results are consistent with previous model results(11). Tabular data for attributable incidence can be found in the Supplementary Material (**Table S2**).

**Figure 1.**
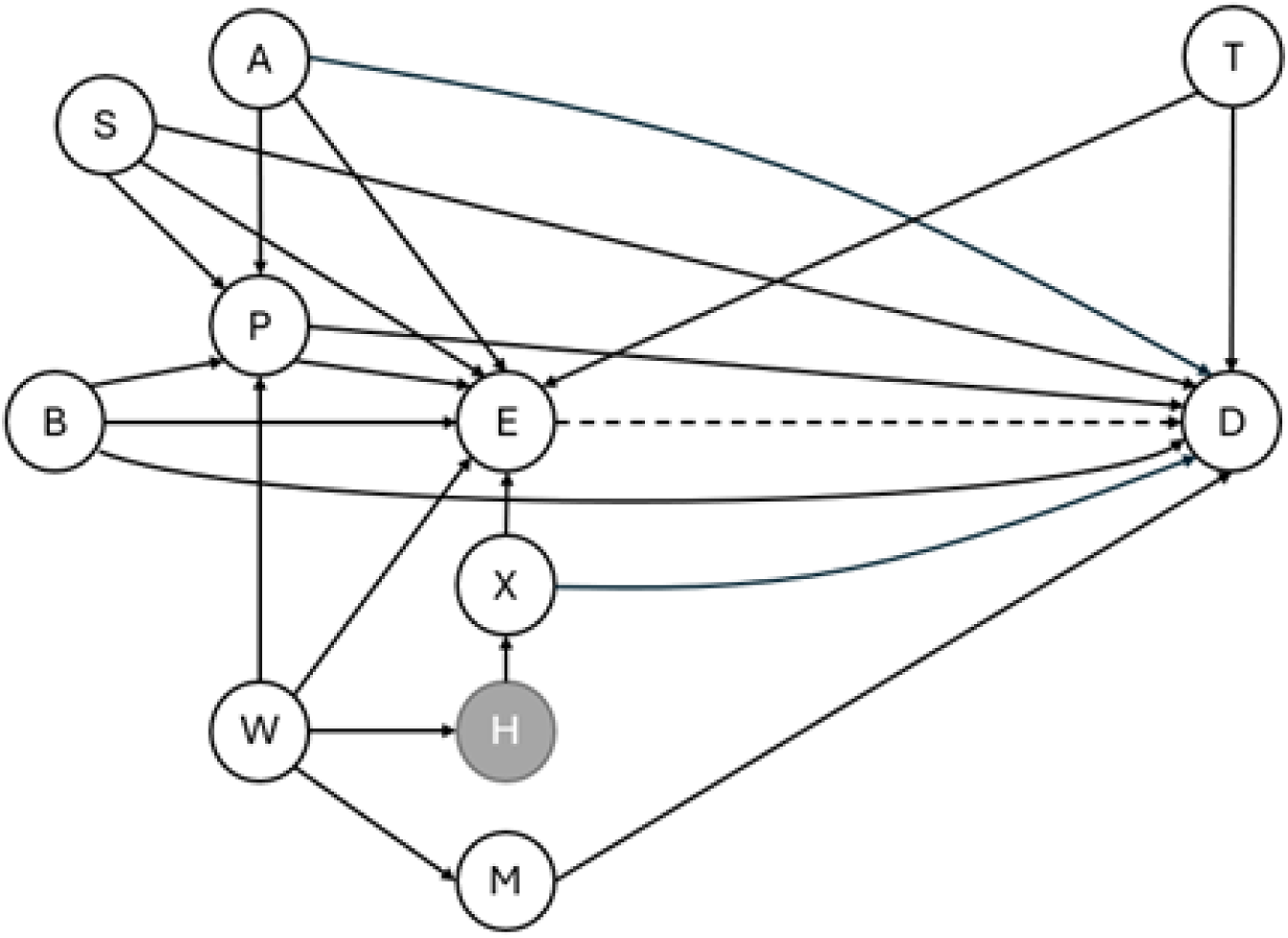
Directed acyclic graph of the association between pathogen quantity and diarrhoea. The causal relationship of interest is shown with a dashed line. Measured covariates are shown in white, and unmeasured covariates are shown in grey. E: pathogen quantity; D: diarrhoea; A: age; B: breastfeeding; H: healthcare resource utilization and literacy; M: malnutrition; P: prior immunizing exposure; S: sex; T: test batch; X: antibiotics use; W: SES, as measured by WAMI score

### Confounder Sensitivity

There was minimal impact of unadjusted confounding on IR_attr_ estimates by antibiotics use prior to the stool sample, prior exposure, breastfeeding, and WAMI score for both the first and second years of life. Adjusting for prior exposure had the greatest impact on IR_attr_ estimates, with adjusted models consistently underestimating attributable incidence compared to the reference model (**Figure 2**). Absolute changes in IR_attr_ point estimates when adjusting for prior infection ranged from 0.2 fewer (norovirus) to 2.0 fewer attributable episodes (sapovirus) per 100 child years in the first two years of life compared to the reference model. The highest relative differences in attributable incidence point estimates generally occurred among the pathogens with the lowest IR_attr_: tEPEC (16% fewer episodes) and cryptosporidium (14% fewer episodes). Attributable incidence estimates for all other pathogens shifted by less than 10% from the reference (**Figure 2**).

**Figure 2.**
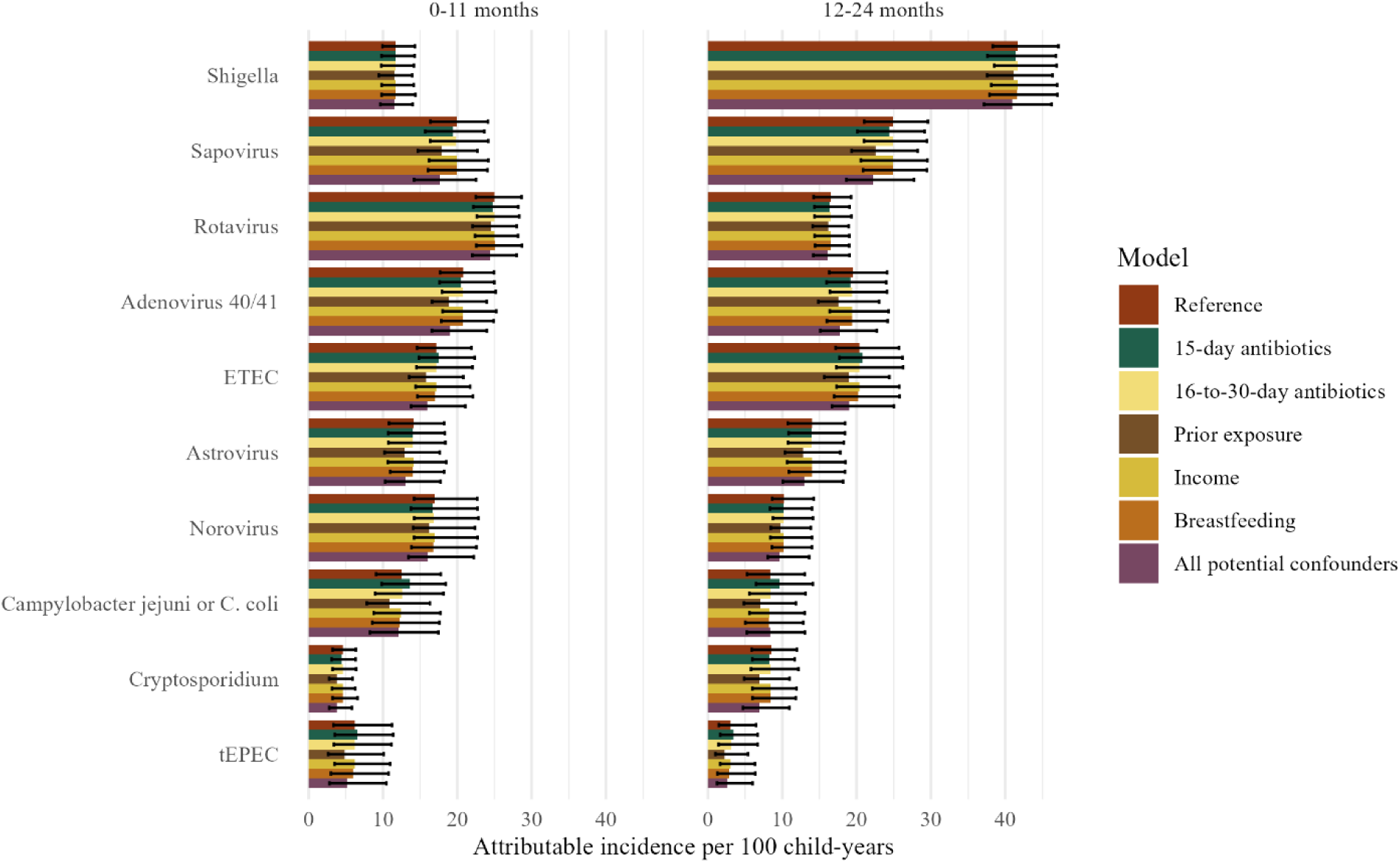
Confounder sensitivity analysis of etiologically-attributable diarrhea incidence over the first two years of life in the MAL-ED cohort, by descending 2-year attributable incidence in the reference model. The reference model is adjusted for quantity of other pathogens, child sex, test batch, child age, a random slope for site, and a random intercept for each child. The legend label corresponds to the covariate added to the reference model. Norovirus includes both GI and GII, and ETEC includes both heat-stable and heat-labile ETEC. Error bars show 95% confidence intervals from 1000 bootstrap estimates.

### Sensitivity on Sample Exclusion

Of 40 406 samples, 3012 occurred ±7 days of a diarrhoeal stool, 6094 occurred ±14 days of a diarrhoeal stool, and 7113 occurred ±28 days of a diarrhoeal stool, resulting in an analytic sample size of 37 394 samples, 34 312 samples, and 33 293 samples for the sensitivity analyses, respectively. Excluding stool samples that occurred near a diarrhoeal stool had little impact on IR_attr_ estimates. Generally, more stringent stool exclusion criteria increased estimates of IR_attr_, but there was no clear trend (**Figure 3**). Over the study period, all point estimates were within 10% of the reference model with the exception of cryptosporidium (7-day: −1.0%, 14-day: +24.4%, 28-day: +25.3%), tEPEC (7-day: 15.6%, 14-day: +29.0%, 28-day: −21.8%), and norovirus (7-day: +8.0%, 14-day: +12.7%, 28-day: +13.9%). All confidence intervals had substantial overlap and included the point estimate from the reference model.

**Figure 3.**
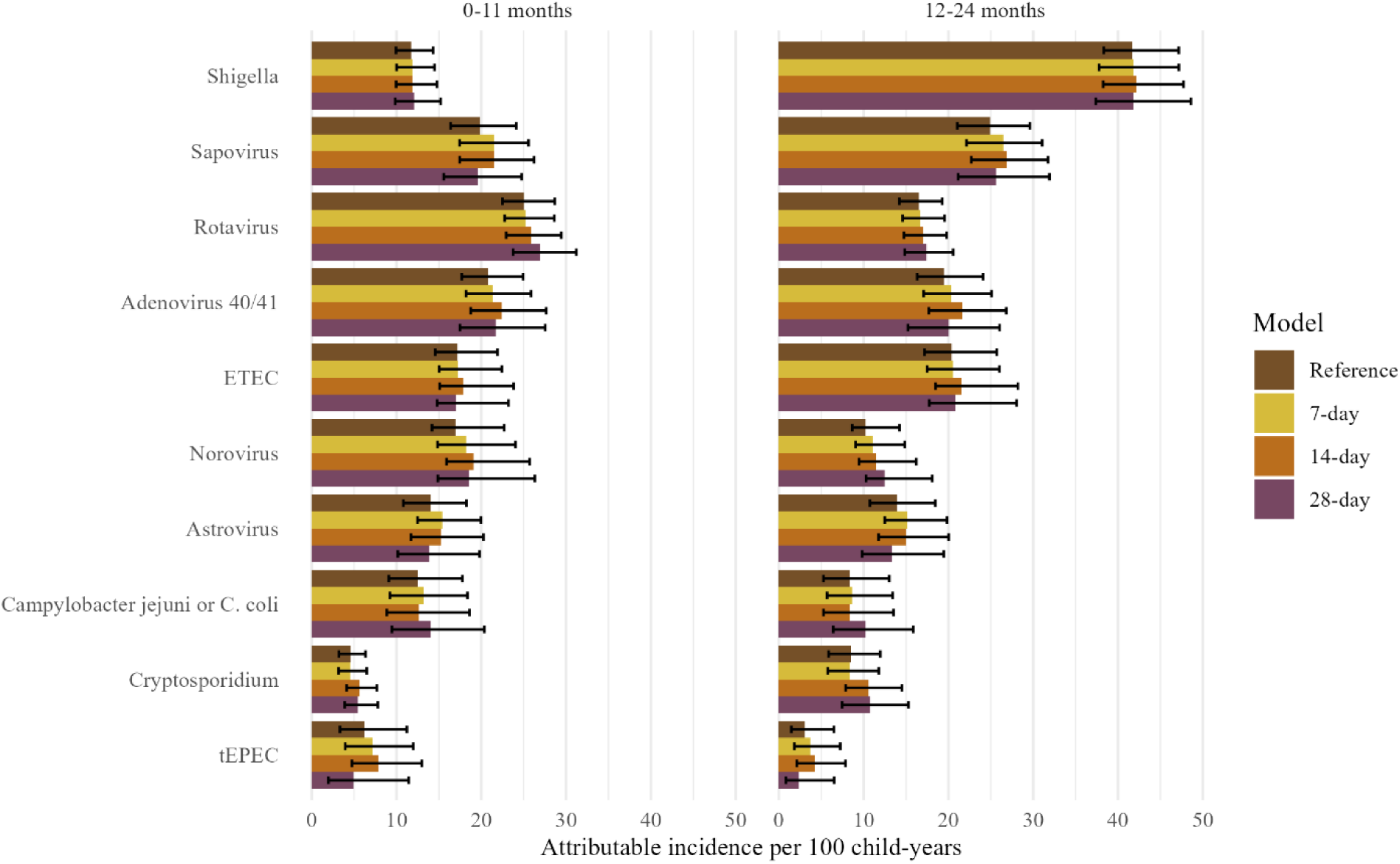
Sensitivity analysis of sample inclusion criteria in the estimation of aetiologically attributable diarrhoea incidence over the first two years of life in the MAL-ED cohort, by descending 2-year attributable incidence in the reference model. The reference model includes all stool samples, where distinct study-defined diarrhoea episodes were separated by at least 48 hours. Sensitivity analyses are conducted excluding all samples that occurred within 7, 14, and 28 days of a diarrheal stool. Error bars show 95% confidence intervals from 1,000 bootstrap estimates.

### Sensitivity on Study Design

Of 6625 case stool samples, 6600 samples (99.6%) from 1258 children were able to be matched to a control stool from a different child for the nested case-control analysis, and 5137 case stools (77.5%) from 1233 children were able to be matched to a control stool from the same child for the nested case-crossover analysis. All samples were included in the risk-based IR_attr_ estimates. In the nested case-control study, the median (Q1, Q3) age difference between matched samples was 0 (−2, 4) days. In the nested case-crossover study, control stools were sampled a median (Q1, Q3) of 23 (13, 31) days prior to the case stool. While there was no consistent pattern of change between the reference model and the matched case-control model, the nested case-crossover model overestimated attributable incidence compared to the matched case-control study design for all pathogens except ETEC. Generally, all odds-based estimates had notable overlap in confidence intervals over the study period (**Figure 4**).

**Figure 4.**
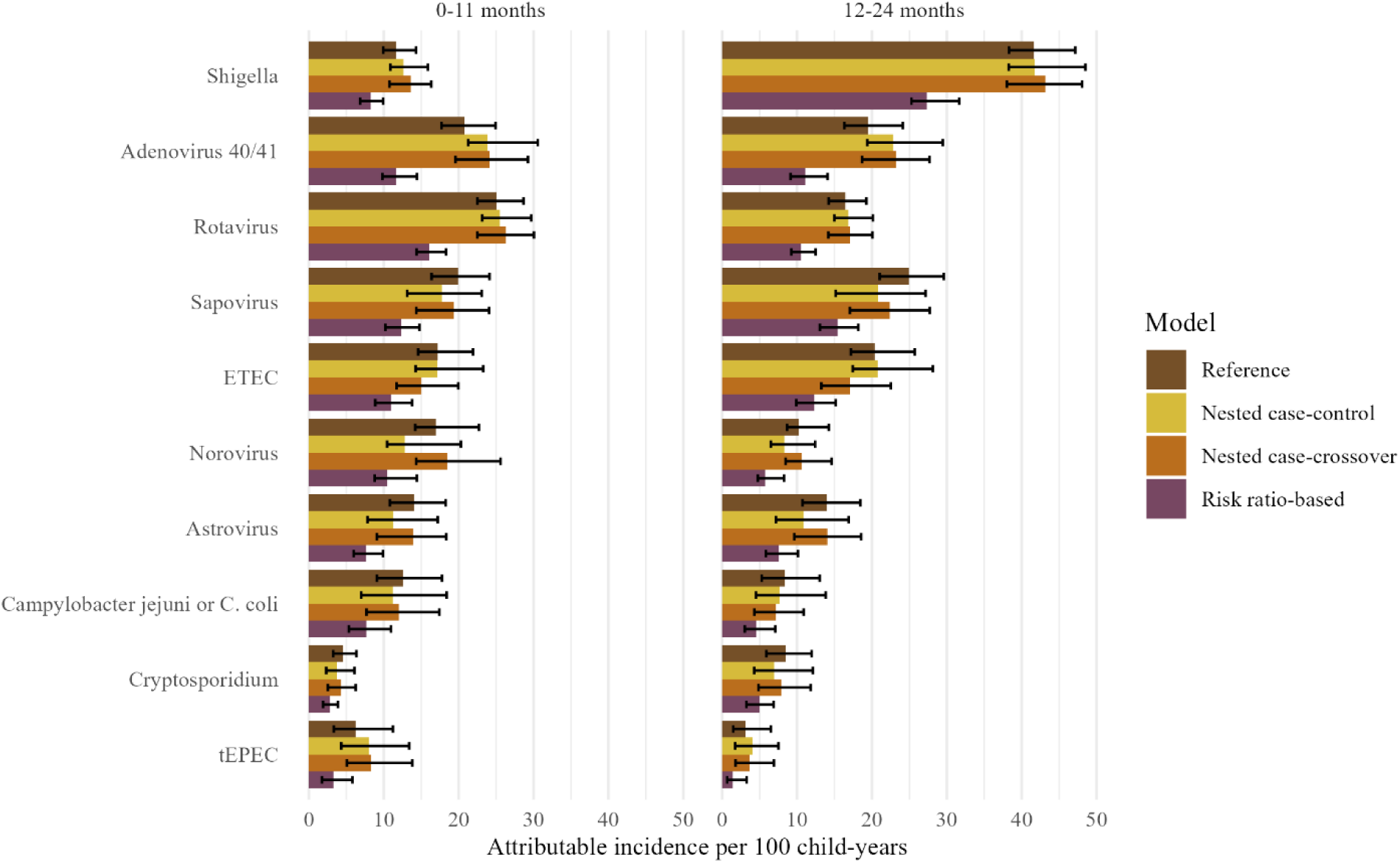
Sensitivity analysis on nested study design in the estimation of aetiologically attributable incidence of diarrhoea in the MAL-ED cohort, by descending 2-year incidence rate in the reference model in the first and second years of life. The nested case-control design used samples matched 1:1 on age within 14 days, sex, and site, and the nested case-crossover design used samples matched on child and age between 4 and 60 days prior to the diarrhoea episode. Error bars show 95% confidence intervals from 1,000 bootstrapped estimates.

Risk-based IR_attr_ estimates were lower than odds-based estimates, although the relative ranking of pathogens was consistent. Relative differences from the reference model ranged from 33% fewer episodes (Shigella) to 49% fewer episodes (tEPEC) per 100 child-years. We observed the largest relative differences in estimates for tEPEC, astrovirus (46% fewer episodes per 100 child-years), and adenovirus 40/41 (44% fewer episodes per 100 child-years).

## Discussion

These sensitivity analyses evaluated the robustness of our previously-developed algorithm to estimate pathogen-specific attributable incidence of diarrhoea in the first two years of life in the MAL-ED cohort. The algorithm was robust to uncontrolled confounding and study-defined diarrhoea. Contrary to our hypothesis that prior exposure would confound estimates downwards, adjusting for prior exposure decreased IR_attr_ estimates across pathogens and there was substantial confidence interval overlap. Diarrhoeal stools were more likely than control stools to have prior immunizing exposure, indicating possible unmeasured confounding by exposure risk or host susceptibility, such that children with prior infections are more likely to be infected again. Adjusting for prior exposure likely partially adjusted for unbalanced exposure risk, but there may be residual confounding still present by unmeasured factors(22). As expected, odds-based models overestimated the incidence of pathogen-attributable diarrhoea compared to risk-based models, but the relative ranking of incidence by pathogen did not change substantially. Mathematically, it is expected that an attributable fraction calculated using a cohort-based odds ratio will overestimate one calculated using a risk ratio when the outcome, in this case diarrhoea, is not rare. In some settings, such as the Global Enteric Multicenter Study (GEMS) which used individually-matched case-control incidence density sampling, the odds ratio will estimate the incidence rate ratio, which in turn is a closer approximation of the risk ratio. Therefore, while odds-based estimates of the IR_attr_ may overstate aetiology in the MAL-ED cohort, they may still provide valid estimates in areas of low diarrhoeal burden or in case-control studies using incidence density or case-base sampling.

Most children experienced at least one infection at a level capable of inducing natural immunity for almost all pathogens, with the exception of norovirus GII and rotavirus, which is consistent with current estimates of diarrhoea prevalence among children in similar settings(1,2). *Shigella*, sapovirus, adenovirus 40/41, and rotavirus were identified as leading causes of diarrhoea, which is consistent with studies conducted in other settings and using differing methods of attribution(6–9). These studies attributed relatively more diarrhoea to ETEC and *Cryptosporidium*, but prolonged post-diarrhoeal shedding may have biased these estimates upwards, and the relative instability of estimates for Cryptosporidium in the sensitivity analysis of study-defined diarrhoea highlights the impact that pathogen shedding can have on attributable incidence estimates. However, this may also be because of lower power to detect associations, especially among low-incidence pathogens, with the smaller sample size as more samples are excluded.

This analysis shows that the AF algorithm is adaptable to many study settings, including case-only designs(26). The MAL-ED cohort provides rich longitudinal data that does not rely on hospital-based case finding and includes testing for a large range of enteropathogens, limiting bias from selection and unmeasured confounding. However, prospective cohorts are often not feasible to study diarrhoeal aetiology. These results show that similar estimates can be obtained from less resource-intensive study designs, such as matched case-control studies, using a small sufficient set of confounders and matching factors: age, geographic location, gender, and a small set of pathogens needed for testing. Additionally, the need to recall prior episodes of diarrhoea is limited, as estimates excluding diarrhoeal stools greater than two days prior were similar to estimates including all episodes.

Despite its longitudinal community-based design, estimating diarrhoeal incidence from the MAL-ED cohort has limitations as previously described(18), including the potential for preferential ascertainment of longer-duration episodes, over-sensitivity of qPCR, and potential unmeasured confounding by exposure or susceptibility. Additionally, breastfeeding and antibiotics use in this analysis were found not to be important confounders, although both have been shown to be associated with pathogen quantity and diarrhoea(14,15). The lack of confounding is therefore unlikely to be transportable to other studies, so it is worth collecting and accounting for recent breastfeeding and antibiotics use in future analyses. Additionally, while minimal, prior immunity from infection had the largest magnitude of confounding bias compared to the reference model. Because prior immunity is difficult to measure in surveillance-based study designs, it is likely impossible to account for in most studies. Finally, this study did not consider cross-immunity between pathogens.

Estimating attributable incidence by the quantity of pathogen detected, rather than assuming all detections are etiologic, is critical in developing reliable estimates of diarrhoeal aetiology. This study shows that the AF-based method for estimation is robust to varying data availability or assumptions about confounding and can be applied in multiple settings. When available, we recommend adjusting for breastfeeding, antibiotics use, and prior immunity. In addition, risk ratio-based algorithms should be used because the rare disease assumption is unlikely to hold in areas where etiological attribution estimates are needed. If a case-control design is necessary, individually-matched incidence density sampling may provide more valid estimates. Results from this analysis add to existing evidence of the key drivers of diarrhoeal burden in high-incidence settings for focus in vaccine development.

## Ethics Approval

All sites received ethics approval from their respective governmental, local institutional, and collaborating institutional ethics review boards. Study staff obtained written consent from the parent or guardian of every child.

## Data Availability

Data for the MAL-ED cohort are publicly available at clinepidb.org.

## Supplementary Data

Supplementary data are available at *IJE* online.

## Author Contributions

CD conducted the data analyses for the present study, drafted the manuscript, and was responsible for manuscript preparation and submission. JPM contributed to data analysis, data curation, development of methodology, and interpretation. JL contributed to methodology, data collection, and data validation. EH led project administration, funding acquisition, and supervision for the parent study. ERM led conceptualization, supervision, and funding acquisition, and contributed to methodology. All authors provided critical review and editing of the draft and provided approval of this version of the manuscript for submission.

## References

1. Institute for Health Metrics and Evaluation (IHME). Global Burden of Disease Study 2021 (GBD 2021) Results [Internet]. 2021 [cited 2025 Jan 15]. Available from: vizhub.healthdata.org/gbd-results

2. Sreeramareddy CT, Low YP, Forsberg BC. Slow progress in diarrhea case management in low and middle income countries: evidence from cross-sectional national surveys, 1985-2012. BMC Pediatr. 2017 Mar 21;17(1):83.

3. Burnett E, Parashar UD, Tate JE. Global Impact of Rotavirus Vaccination on Diarrhea Hospitalizations and Deaths Among Children <5 Years Old: 2006-2019. J Infect Dis. 2020 Oct 13;222(10):1731–9.

4. Armah G, Lopman BA, Vinjé J, O’Ryan M, Lanata CF, Groome M, et al. Vaccine value profile for norovirus. Vaccine. 2023 Nov 3;41 Suppl 2(Suppl 2):S134–52.

5. MacLennan CA, Grow S, Ma LF, Steele AD. The Shigella Vaccines Pipeline. Vaccines. 2022 Aug 24;10(9):1376.

6. Thystrup C, Majowicz SE, Kitila DB, Desta BN, Fayemi OE, Ayolabi CI, et al. Etiology-specific incidence and mortality of diarrheal diseases in the African region: a systematic review and meta-analysis. BMC Public Health. 2024 Jul 12;24(1):1864.

7. Gasparinho C, Mirante MC, Centeno-Lima S, Istrate C, Mayer AC, Tavira L, et al. Etiology of Diarrhea in Children Younger Than 5 Years Attending the Bengo General Hospital in Angola. Pediatr Infect Dis J. 2016 Feb;35(2):e28–34.

8. Huilan S, Zhen LG, Mathan MM, Mathew MM, Olarte J, Espejo R, et al. Etiology of acute diarrhoea among children in developing countries: a multicentre study in five countries. Bull World Health Organ. 1991;69(5):549–55.

9. Vu Nguyen T, Le Van P, Le Huy C, Nguyen Gia K, Weintraub A. Etiology and epidemiology of diarrhea in children in Hanoi, Vietnam. Int J Infect Dis. 2006 Jul;10(4):298–308.

10. McMurry TL, McQuade ETR, Liu J, Kang G, Kosek MN, Lima AAM, et al. Duration of Postdiarrheal Enteric Pathogen Carriage in Young Children in Low-resource Settings. Clin Infect Dis Off Publ Infect Dis Soc Am. 2021 Jun 1;72(11):e806–14.

11. Platts-Mills JA, Liu J, Rogawski ET, Kabir F, Lertsethtakarn P, Siguas M, et al. Use of quantitative molecular diagnostic methods to assess the aetiology, burden, and clinical characteristics of diarrhoea in children in low-resource settings: a reanalysis of the MAL-ED cohort study. Lancet Glob Health. 2018 Dec;6(12):e1309–18.

12. Liu J, Platts-Mills JA, Juma J, Kabir F, Nkeze J, Okoi C, et al. Use of quantitative molecular diagnostic methods to identify causes of diarrhoea in children: a reanalysis of the GEMS case-control study. The Lancet. 2016 Sep;388(10051):1291–301.

13. McCormick BJ, Richard SA, Murray-Kolb LE, Kang G, Lima AA, Mduma E, et al. Full breastfeeding protection against common enteric bacteria and viruses: results from the MAL-ED cohort study. Am J Clin Nutr. 2022 Mar;115(3):759–69.

14. Turin CG, Ochoa TJ. The Role of Maternal Breast Milk in Preventing Infantile Diarrhea in the Developing World. Curr Trop Med Rep [Internet]. 2014 Mar 15 [cited 2025 Feb 7]; Available from: http://link.springer.com/10.1007/s40475-014-0015-x

15. Long K, Vasquez-Garibay E, Mathewson J, de la Cabada J, DuPont H. The impact of infant feeding patterns on infection and diarrheal disease due to enterotoxigenic Escherichia coli. Salud Publica Mex. 1999;41(4):263–70.

16. The MAL-ED Network Investigators, Acosta AM, Chavez CB, Flores JT, Olotegui MP, Pinedo SR, et al. The MAL-ED Study: A Multinational and Multidisciplinary Approach to Understand the Relationship Between Enteric Pathogens, Malnutrition, Gut Physiology, Physical Growth, Cognitive Development, and Immune Responses in Infants and Children Up to 2 Years of Age in Resource-Poor Environments. Clin Infect Dis. 2014 Nov 1;59(suppl 4):S193–206.

17. Houpt E, Gratz J, Kosek M, Zaidi AKM, Qureshi S, Kang G, et al. Microbiologic Methods Utilized in the MAL-ED Cohort Study. Clin Infect Dis. 2014 Nov 1;59(suppl_4):S225–32.

18. Platts-Mills JA, Babji S, Bodhidatta L, Gratz J, Haque R, Havt A, et al. Pathogen-specific burdens of community diarrhoea in developing countries: a multisite birth cohort study (MAL-ED). Lancet Glob Health. 2015 Sep;3(9):e564–75.

19. Bruzzi P, Green SB, Byar DP, Brinton LA, Schairer C. Estimating the population attributable risk for multiple risk factors using case-control data. Am J Epidemiol. 1985 Nov;122(5):904–14.

20. Lewnard JA, Lopman BA, Parashar UD, Bar-Zeev N, Samuel P, Guerrero ML, et al. Naturally Acquired Immunity Against Rotavirus Infection and Gastroenteritis in Children: Paired Reanalyses of Birth Cohort Studies. J Infect Dis. 2017 Aug 1;216(3):317–26.

21. Lewnard JA, Lopman BA, Parashar UD, Bennett A, Bar-Zeev N, Cunliffe NA, et al. Heterogeneous susceptibility to rotavirus infection and gastroenteritis in two birth cohort studies: Parameter estimation and epidemiological implications. Viboud C, editor. PLOS Comput Biol. 2019 Jul 26;15(7):e1007014.

22. Rogawski McQuade ET, Liu J, Kang G, Kosek MN, Lima AAM, Bessong PO, et al. Protection From Natural Immunity Against Enteric Infections and Etiology-Specific Diarrhea in a Longitudinal Birth Cohort. J Infect Dis. 2020 Nov 9;222(11):1858–68.

23. Psaki SR, Seidman JC, Miller M, Gottlieb M, Bhutta ZA, Ahmed T, et al. Measuring socioeconomic status in multicountry studies: results from the eight-country MAL-ED study. Popul Health Metr. 2014 Dec;12(1):8.

24. Garcia Quesada M, Platts-Mills JA, Liu J, Houpt ER, Rogawski McQuade ET. Leveraging Data From a Longitudinal Birth Cohort to Improve Attribution of Diarrhea Etiology Among Children in Low-Resource Settings. J Infect Dis. 2024 Aug 7;jiae389.

25. Qiu Y, Freedman SB, Williamson-Urquhart S, Farion KJ, Gouin S, Poonai N, et al. Significantly Longer Shedding of Norovirus Compared to Rotavirus and Adenovirus in Children with Acute Gastroenteritis. Viruses. 2023 Jul 13;15(7):1541.

26. Cohen AL, Platts-Mills JA, Nakamura T, Operario DJ, Antoni S, Mwenda JM, et al. Aetiology and incidence of diarrhoea requiring hospitalisation in children under 5 years of age in 28 low-income and middle-income countries: findings from the Global Pediatric Diarrhea Surveillance network. BMJ Glob Health. 2022 Sep;7(9):e009548.

